# Giving patients a voice within healthcare reform: the qualitative VOICE study

**DOI:** 10.1101/2021.05.11.21256986

**Authors:** Kathryn Hoffmann, Nicole Rumpler, Aaron George, Pauline de Boeckxstaens

## Abstract

**Background:** Inclusion of patients in healthcare service and system planning is an increasingly important tool to improve healthcare systems worldwide. In 2012, a focused healthcare reform was initiated in Austria to strengthen the primary care sector.

**Objectives:** The aim of this study was to assess the perceptions, desires and needs of patients in terms of primary care as a necessary building block of the Austrian healthcare reform.

**Methods:** This study was designed as an exploratory qualitative study using semi-structured interviews between the years 2013 and 2018. Research questions focused on patients’ positive and negative experiences with regard to GP consultations and the overall primary care system, as well as desires for improvement. Qualitative content analysis by Mayring was used to analyze the material.

**Results:** Altogether, 41 interviews were conducted with seven main-categories identified. These categories include coordination and time management around consultation, human and professional aspects of consultation including coordination of care, access and availability including opening hours, infrastructure and hygiene of the waiting room, personnel, and overarching healthcare system factors.

**Conclusion:** This study demonstrates the importance of bringing the patients’ voice into ongoing healthcare reform. Without appreciating and responding to patients’ perceptions and needs, healthcare reform in Austria might be challenged to improve in areas such as time, coordination and navigation. Successful health care reform necessitates the inclusion of the patient voice.

## Introduction

Inclusion of patients in healthcare service and system planning is an increasingly important tool to improve healthcare systems worldwide (1-4). Decades of research supports that patient satisfaction is tied to availability, accessibility, accommodation, acceptability, and affordability of care (5, 6). In the U.K., for example, the involvement of patients is central to promotion of continuous improvement in quality of care delivery (3). Further, a global perspective of care delivered is more inclusive and offers potential to be more responsive. This is particularly relevant for those who have financial investment in the healthcare system through their social health insurance companies’
s fees and taxes (4).

Bochel et al. stated in their review of participation that *“participation might be intended to improve governance, democracy, social capital, education and development of individuals, policies, service implementation and delivery (1)*.*”* In terms of healthcare planning and development, this suggests that patient involvement could lead to improved development of services, structures and outcomes of care (2, 3).

In 2012, Kringos et al. published a revealing document on the strength of primary care in Europe, and within this Austria was identified with relatively underdeveloped primary care (7, 8). Austria was found to have weaknesses in the governance of primary care (PC), workforce development, and coordination and continuity of care (7, 8). Historically, the public Austrian healthcare system has been structured with two levels of care, which include the hospital and ambulatory sectors. This is disparate from traditional models elsewhere which typically involve three levels of care (primary, secondary and tertiary level) (9). This structure is a barrier to the continuity and coordination role of primary care. Additionally, Austria has universal health coverage, but no clear demarcation line for access between primary and secondary care (9). Individuals can access with few exceptions secondary specialist care in the ambulatory sector without knowledge or facilitation of a GP, and they do so in much higher numbers than the rest of Europe (9, 10). In addition, mainly solo GP practices exist and number of primary care teams across the country remains low (11). Consequently, a healthcare reform has been initiated in Austria, with the aim to strengthen the primary care sector.

In a first step, Austrian primary care reform aims to facilitate and increase the number of GP group practices with primary care teams, expand opening hours, and enhanced service profiles either via centres or via networks (11). This approach aims to make PC practices more comprehensive and attractive for patients to access them as an initial point of care without having to add a clear demarcation line between primary and secondary care on a structural level. The first primary care centre was established in 2015 to encourage such access. As of 2021, with reform ongoing, there are now 24 primary care centres and networks established (12).

Alongside system and infrastructure development, it is essential to involve the voices of those receiving care. Given this imperative, the aim of this study was to understand what patients value and identify for improvement in their perception of the existing publicly funded primary care setting by collection and analysis of people’s experience of GP consultations.

## Methods

This study was designed as an exploratory qualitative study using semi-structured interviews in accordance with the COREQ checklist. As qualitative research aims to gain deeper insight into a topic and understand the perception of persons beyond prepopulated questionnaires, it is an ideal method for such research wherein understanding of local context and perception is limited.

### Recruitment

The recruitment of participants took place in the three most populous counties of Austria, namely Vienna, Lower Austria, and Upper Austria, via the snowball system. Vienna is the capital and is additionally a county as well as the only metropolis in Austria. Lower Austria is a more rural county and Upper Austria is mixed with industrial sites and rural areas. Four medical students used identical recruitment techniques and interview guidelines to conduct the interviews after being throughout trained in interview technique (13-16). In Vienna, one student recruited German speaking persons and the other Turkish speaking persons because Turkish individuals represent the third largest group of migrants in Vienna, after Serbs and Germans, which speak the same language as Austrians (17). For the snowball system, each student recruited two persons within their circle of acquaintances. After finishing the interviews, these persons recommended two new participants in a consistent manner until data saturation was reached for each county. The aim was to recruit equal numbers of women and men, as well as equal persons from five age groups, namely 18-30, 31-45, 46-60, 61-75 and 76 years and older. Other inclusion criteria were compos mentis, living in the county for a minimum of five years and having visited a publicly funded GP within the county at least once. All participants were informed about the study prior to their participation and had to sign an informed consent if they were willing to participate. None of the interview partners dropped-out during the interview.

### Data collection

The participants were given the option to interview in an office at the Medical University of Vienna or at their homes. The setting for the interviews was a one-to-one between only the interviewer and the interviewee. Before the start of the interview, the interviewees stated their sex, age group, and highest educational level in a short questionnaire, which was linked to the interview via a specific code. Semi-structured interviews were conducted for all participants. The interviews took between 30 and 90 minutes, with a mean of 60 minutes, and were digitally recorded. None of the interviews needed to be repeated.

The interview guideline for the semi-structured interviews contained five questions on their needs and desires for improving their primary care experience.

1. Tell me about your last GP consultation at a public GP practice
2. What do you think about primary care in your county in general?
3. What do you like regarding your healthcare provided by GPs and GP practices? Why? Do you have examples?
4. What do you not like regarding your healthcare provided by GPs and GP practices? Why? Do you have examples?
5. If you would have the opportunity what would you like to change and how?

Additional questions were raised on satisfaction and utilization of healthcare services if participants were not immediately responsive (5, 6).

### Data analysis

All digitally recorded interviews were transcribed, with the Turkish transcripts additionally translated into German. Content analysis was performed under the framework of Mayring (18).

The second author (NR) who was not involved in the interviews analysed all original transcripts, in accordance with the analysis steps defined by Mayring (18). A summarizing inductive procedure was chosen in which a category was formed according to *“the most naturalistic, object-oriented depiction of the material without distortion due to the researcher’s assumptions”* (18) and, thus, reflects the *“language of the material”* (18). Secondarily, for those areas demonstrating need of improvement were assigned deductively based on the structure of the interview guideline. The first author (KH) coded at least 30% additionally, codes were compared and discussed by the two authors to check for coder reliability in terms of each of the main categories. Thus, deeper meaning of the material could be analysed and phenomena could be described.

In addition, the qualitative content analysis by Mayring as a hypothesis-generating method offers the advantage that, through the possibility of integrating frequency counts of categories and sub-categories, qualitative results can also be quantified (18). Frequencies are thus interpreted with caution in such a way, that the more often a category is mentioned by the participant, the more important, obvious, and significant it might be for them.

The mentioning of a sub-category was counted as one when it was given at least once in an interview; and the same was performed for main categories.

For the purpose of this publication, the quotes were translated into the English language by the authors NR and KH.

### Ethical considerations

This study had a positive ethical approval by the ethics committee of the Medical University of Vienna (Nr. 224/2019) and were conducted under European law regarding data protection and security.

## Results

Overall, 41 interviews could be conducted. Table 1 shows the demographics of the participants.

**Table 1:** Demographics of the participants

| Variable | Sub-variable | n | % |
| --- | --- | --- | --- |
| Overall |  | 41 | 100 |
| Sex | female | 18 | 43,9 |
|  | male | 23 | 56,1 |
| Age group | 18 - 30 years | 10 | 24,4 |
|  | 31 – 45 years | 7 | 17,1 |
|  | 46 – 60 years | 13 | 31,7 |
|  | 61 – 75 years | 7 | 17,1 |
|  | 76 years and older | 4 | 9,8 |
| Highest completed educational level | primary | 8 | 19,5 |
|  | secondary | 20 | 48,8 |
|  | tertiary | 13 | 31,7 |
| County | Vienna – German speaking | 10 | 24,4 |
|  | Vienna – Turkish speaking | 8 | 19,5 |
|  | Lower Austria | 12 | 26,8 |
|  | Upper Austria | 11 | 29,3 |

### Description of the analysed main- and sub-categories

Overall, 711 statements were made by the forty-one interviewees. 7 main categories and 39 related sub-categories could be analysed from these statements. The initial main category contains aspects of access, availability, and opening-hours of GP practices, the second contains organisational aspects around consultation, and the third on accommodation and infrastructure. The fourth and fifth categories each contain two different aspects regarding the treatment itself, namely professional and human aspects of care. The sixth category subsumes aspects regarding the personnel within the GP practices, and the seventh is related to superordinate healthcare factors.

Table 2 gives an overview of all main- and related sub-categories as well as if a category and sub-category was mentioned at least once in an interview.

**Table 2:** Main and sub-categories as well as frequency of mentions.

| Main category | % (n) of mentions within the main categories (N=41) | Sub-category | % (n) of mentions of the sub-categories within specific main category |
| --- | --- | --- | --- |
| Access | 87,8 (36) | Opening-hours | 55,6 (20) |
|  |  | Availability of GP services in the vicinity | 41,7 (15) |
|  |  | Accessibility | 13,9 (5) |
|  |  | Consultations via telemedicine | 11,1 (4) |
|  |  | Home visits | 30,6 (11) |
|  |  | Emergency service | 8,3 (3) |
| Organisation/time management around the consultations | 100,0 (41) | Appointment service | 65,9 (27) |
|  |  | Waiting time and circumstances prior to consultation | 85,4 (35) |
|  |  | Number of patients in the waiting room | 35,0 (14) |
|  |  | Duration of the consultation/consultation length | 53,7 (22) |
| Accommodation/ infrastructure of the GP practice | 78,0 (32) | Technical infrastructure (ECG, point-of-care ultrasound, small laboratory etc.) | 34,4 (11) |
|  |  | Adequate number and kind of chairs in the waiting room | 31,3 (10) |
|  |  | Size and ambience of the waiting room | 28,1 (9) |
|  |  | Availability of information material/information via TV/water in the waiting room | 50,0 (16) |
|  |  | Hygiene, air circulation and light | 25,0 (8) |
|  |  | Privacy at the reception desk | 15,6 (5) |
|  |  | Special waiting room/area for children with parents | 28,1 (9) |
| Consultation (human aspects) | 87,8 (36) | Established relationship | 58,3 (21) |
|  |  | Trust and reliability | 13,9 (5) |
|  |  | Interest in the patients living conditions | 33,3 (12) |
|  |  | Empathy and being taken seriously | 52,8 (19) |
|  |  | Caring behaviour of the GP | 22,2 (8) |
|  |  | Characteristics of GP and patient fit | 33,3 (12) |
|  |  | Stress of the GP | 22,2 (8) |
| Consultation (professional aspects) | 97,6 (40) | Professional communication skills including active listening | 70,0 (28) |
|  |  | Coordination and navigation function | 70,0 (28) |
|  |  | Up to date diagnostic and therapeutic skills, accuracy | 67,5 (27) |
|  |  | Up to date (continuous) medical education | 47,5 (19) |
|  |  | Prescription of medications | 57,5 (23) |
|  |  | Broadening of services or additional services offered (e.g. additional PC professions in the practice, preventive services, alternative medicine) | 60,0 (24) |
| Personnel | 41,5 (17) | Manner of dealing with patients | 88,2 (15) |
|  |  | Competency | 29,4 (5) |
| Overarching healthcare system factors | 78,0 (32) | Bureaucracy due to regulations of social health insurance companies | 40,6 (13) |
|  |  | Payment of GPs | 31,3 (10) |
|  |  | Services that are and are not financially supported by social health insurance companies | 37,5 (12) |
|  |  | Supportive of GP group practices | 37,5 (12) |
|  |  | Regulations for success of GP practices | 12,5 (4) |
|  |  | Superordinate quality management | 18,8 (6) |
|  |  | Free choice of GP | 25,0 (8) |

In terms of positive, negative and improvable aspects of the categories and sub-categories, a causal directionality was prominent for all categories: positive related to appreciation of an existing aspect of the category, negative to one that was missing and improvable for one to be adapted.

In depth analysis showed that the category *“organisation and time management around the consultation”* was found to be of importance for participants, in particular with respect to the *“waiting time and circumstances before the consultation”*. Respondents preferred the option to make an appointment ahead of time, as opposed to presenting to a GP practice and waiting. Another important factor is the importance of appropriate time and duration of consultation.

> *“I could visit him anytime, but I knew I had to wait 3-4 hours if I went there without an appointment. Or if I was sick, my health would even worsen. For example, when I was there because my arm was sore I waited 3 hours. And a week later I was there again with the flu because I got infected there*.*”* (Participant 31)

With regard to the *“professional aspect of the consultation”*, all sub-categories were found to be equal in terms of importance to participants. “Prescribing medications”, however, was one sub-categories frequently mentioned in a negative way, in particular, when it came together with perceived short consultation time.

> *“They prescribe medication relatively quickly, pain killers, sometimes simply without any diagnosis. That is really fatal in some cases*.*”* (Participant 2)

When it comes to the coordination function of GPs regarding patient navigation through the healthcare system, participants desired improvement of this role, as they vocalized being increasingly overwhelmed with the complex pathways of the healthcare system.

> *“I had to organize everything alone. And to call the GP and ask him: “Where shall I go now?” Better to call immediately the ambulance*.*”*
>
> — (Participant 12)

Three points mentioned frequently when talking about *“broadening of services or additional services”* were the desires for improvement of preventive services, and for additional services such as psychologists, physiotherapists, dieticians, occupational therapists under the same roof, and for alternative medicine.

> *“I think that the additional services don’t need to be provided by general practitioners. But GPs should know about alternative therapies, beside prescribing medication, about physiotherapy, whatever. He should know about massage techniques, which could help, and, thus, offer alternative treatments*.*”*
>
> — (Participant 2)

The category “*human aspects of consultation”* was considered as a very important corner stone of a successful consuultation, in particular the factors *“established relationship”, “empathy of the GP”*, and the *“GP’
ss interest in the living conditions”*.

> *“With the physician where I am now, everything is perfect. She takes care for me. She already knows me that well that she can already see on my face how I am feeling. She also responds to my psychological needs, what I really like. I can also visit her without any complaint, just to talk with her, that really impresses me*.*”*
>
> — (Participant 1)

The factor of *“opening-hours”* seems to be the most relevant factor regarding *“access”* to primary care. In terms of accommodation aspects, the infrastructure of the waiting room plays a major role but only if waiting times are long. Under these circumstances participants expressed interest in the availability of information via flyers or TV, of water and adequate seating. Similarly, important is the overall cleanliness of the practice and regular circulation of fresh air.

The category *“overarching healthcare system factors”* were found unexpectedly. Participants mentioned quite detailed measures on overarching levels of the healthcare system to improve the situation at the practice level. For example, social health insurance companies were identified for need for improvement, and this included bureaucratic barriers, electronic prescriptions, referrals, and sick leave certificates. Moreover, GPs should be paid differently and group practices should be enabled and supported.

> *“Because if there are many health professions at the same location it is easier for patients to get all the services needed under one roof. It is easier and more comfortable and probably also more efficient, because the practitioner can communicate also with the others with help of information technology. That should be kept and even expanded*.*”*
>
> — (Participant 5)

> *“I think that from an organizational point of view it would be important to pay medical services in a different way. Not only – medical consultation, paying fee-for-service. Instead of that financing should simply work differently. That mass of persons in the practice – I don’t know how to say – but mass shouldn’t be necessary to finance good medical practice. Instead of that quality should be rewarded because like this patients receive better treatment*.*”*
>
> — (Participant 3)

### Differences regarding the categories and participant’ ss demographics

No differences of the frequencies with regard to the method by Mayring as well as regarding the in depth analysis results between sex groups, age groups and county groups could be found for all seven main categories.

## Discussion

When patients in Austria are given a voice about their perceptions and desires regarding their healthcare by GP practices, they talk about seven areas of importance. These areas are coordination and time management around consultation, professional and human aspects within consultation, access to care including availability and opening hours, infrastructure and hygiene of the waiting room, personnel, and overarching healthcare system factors (table 2).

It is not surprising that the availability of PC services within their vicinity as well as adequate opening hours are important factors for the participants, as these are foundational elements to access to care (5-7). Adequate opening hours are particularly important for those individuals whose employment limits opportunity for seeking daytime care. Morgan et al. showed that satisfaction with opening-hours improved slightly for practices offering extra appointments, for example on Saturdays (19). Additionally, an extension of the daily coverage of primary care services might reduce inappropriate admissions to emergency services (20).

Most important for the participants, however, is the category of organisation and time management around consultation and, in particular, the waiting time before the consultation in the waiting room (table 2). Previous publications have shown that waiting time is heavily associated with satisfaction with the service, as well as willingness to return (21, 22). Long waiting times increase exposure to patients within a shared waiting space, and have also been shown to increase risk of infection (23).

This factor is of additional relevance with regard to the ongoing COVID19 pandemic. In this context, also the infrastructure of the practice and in particular of the waiting room is important. Participants in this study mentioned the importance of modern technical equipment as well as a desire for visible hygiene and fresh air. Moreover, adequate seating and the availability of information is a frequent mentioned desire of participants (table 2).

Another important factor found in this study regarding time management is the length of the consultation itself. Previous publications have shown that time spent with the physician was the strongest predictor of patient satisfaction (21). In addition, this factor is closely associated with waiting time. “*The decrement in satisfaction associated with long waiting times is substantially reduced with increased time spent with the physician [*…*] (21)*. The consultation time in GP practices in Austria is approximately 5 minutes, which is in stark contrast to e.g. Sweden which as a country has a mean time of 22 minutes (24). Five minutes is a remarkably short period to address all the desired professional and human factors within the consultation that are relevant for the study participants (table 2). In addition, consultation length has implications for patient safety, management of chronic disease, diagnoses and outcomes (24, 25).

One study by Odgen et al. shows that the majority of patients might underestimate consultation length. In this study a preference for more time was correlated with a dissatisfaction of the emotional (human) aspects of the consultation but not with the information and examination components of the consultation (26). This shows the relevance of human factors for forming a successful doctor-patient relationship. The doctor-patient relationship influences patients’ compliance with treatment and is an important cornerstone for continuity of care. *“Satisfaction with the doctor–patient relationship is a critical factor in people’s decisions to join and stay with a specific organization (27)*.*”* In Austria, these factors persist, and this is in part due to free choice of GPs, an element of the system that must be maintained (28).

The second most frequently identified category was the *“professional aspects of a consultation”*. In this category, all sub-categories were found to be similarly important to participants (table 2). It is thus essential for GPs to have proper and up-to-date communication skills including the competency of active listening and that they have up-to-date diagnostic and therapeutic skills. In particular, the prescribing of medications should follow from proper diagnosis with counselling. Participants offered concern with adherence following prescriptions done too fast or of too high a quantity or dosage. Another desire expressed in this study was to expand or supplement the existing services by including supplementary primary care professions into the GP practice. This desire is echoed by international publication demonstrating methods to improve patient care (29, 30).

In addition, participants of this study urgently called for a better coordination und navigation function between the level of care and within primary care. As described above in detail, in Austria, first contact and coordination with primary care are not required (9, 10). In contrast, *“[a] strongly developed primary care sector, as defined by the expert panel on effective ways of investing in health (EXPH), commissioned by the European Commission, should ‘play a central role in the overall coordination and continuity of people’s care’ (29)*. Such a coordination and navigation role could protect patients from unnecessary harm due to getting lost in the complex healthcare system, avoiding unnecessary and multiple diagnostics, unnecessary hospital admissions, or wrong treatment strategies (8, 29, 30).

### What are the implications for primary care reform and reform in Austria?

Fortunately, several aspects of Austrian healthcare reform goals align with those perceptions and desires expressed in this study. These include expanded office hours, expanded and enhanced service profiles, integration of an interdisciplinary team into practices, and support of group practices and primary care centres. In addition, free choice of GPs and universal health coverage should be maintained.

However, other important factors relevant to patients’ satisfaction and willingness to access and return to a service remain in need of improvement. These are the organisation and time management around consultation, consultation length that is necessary for all desired and necessary human and professional aspects of consultation, and the coordination function of PC.

In this study, overarching system factors were additionally identified as “*respondents were quick to note that existing fee-for-service payment does not reimburse care coordination efforts. Because there are no payments for such activities as following up on referrals or communicating with patients outside of the office, physicians do so at the expense of other, billable activities (31)*.*”* Moreover, a primarily fee-for-service payment schema, as exists in Austria, correlates strongly with more patients and longer waiting times in a practice. This is in contrast to value- and need-based payment schemes (32).

Most participants were aware that their suggestions for improving primary care generally lie outside of the hands of the GPs and the local practices themselves. Rather, that larger systemic factors, policies, and priorities of health insurance companies drive much of the way care is accessed and delivered. Interestingly, those factors acknowledged by the participants of this study were also identified as critical by Kringos in 2012 and have been recommended at the international level for comprehensive and effective primary care (7, 8, 29, 30).

### Strengths and limitations

One strength of this study is the fact that it is the first of its kind in Austria. Another strength is the approach, employing an exploratory and qualitative method.

The sample of participants is both a strength and a limitation. With 41 participants from different age groups, sex, counties, and language backgrounds, data saturation could easily be reached for this sample. However, with regard to age and educational level, more individuals with higher educational levels from the age group 46 until 60 years of age were included. Additionally, no participants from the more rural southern and western parts of Austria were recruited, potentially missing aspects from these regions.

Another limitation is that, though the qualitative content analysis by Mayring (18) has the advantage to underline results with frequencies, the frequency of categories does not always correlate with their importance. Furthermore, issues such as memory bias, recall bias or bias of possible social desirability could have had an impact on reports of interviewees in this study.

As a last limitation, it must be mentioned that the snowball sampling method bears risk of a community bias, where the first contact has a dominant impact on sample and represents a potentially limited network.

## Conclusion

This study shows the importance of bringing the patient voice into ongoing healthcare reform.

Appreciating and acknowledging the perception of patients provides deeper context and perspective into healthcare access and delivery. New aspects could be found like the desire for a coordination and navigation function for general practice, inter-professional care teams as well as improved time aspects around the consultation. As time and coordination aspects of primary care are improved, structural changes will be necessary alongside those for full system reform and improvement.

For the next steps of reform patients’ voices should be integrated on a regular base to evaluate the success and the achievement of the objectives of reform.

## Supporting information

COREQ checklist

## Data Availability

All data (transcripts) are available upon request at the first and corresponding author.

## Acknowledgments

We would like to thank the four medical students (Georg Förster, Sümeyye Kaya, Paul Amanshauser, Jonathan A. Kraml) for data collection and transcripts as well as Gerald Bachinger, Susanne Rabady, Sebastian Huter, David Wachabauer, Sarah Burgmann, Herwig Ostermann and Vera Krambeer for their valuable input for the discussion section.

## Financial resources

This project was partly funded by the Medical Scientific Fund of the Mayor of the City of Vienna under the number 20032.

## References

1. Bochel C, Bochel H, Somerville P, Worley CJSP, Society. Marginalised or enabled voices?’User participation’in policy and practice. 2008;7(2):201–10.

2. Crawford MJ, Rutter D, Manley C, Weaver T, Bhui K, Fulop N, et al. Systematic review of involving patients in the planning and development of health care. BMJ. 2002;325(7375):1263.

3. NHS Future Forum. Patient involvement and public accountability : a report from the NHS Future Forum. London: 2011.

4. Gagliardi AR, Lemieux-Charles L, Brown AD, Sullivan T, Goel V. Barriers to patient involvement in health service planning and evaluation: An exploratory study. Patient Education and Counseling. 2008;70(2):234–41.

5. McLaughlin CG, Wyszewianski L. Access to care: remembering old lessons. Health services research. 2002;37(6):1441–3.

6. Penchansky R, Thomas JW. The Concept of Access: Definition and Relationship to Consumer Satisfaction. Medical Care. 1981;19(2):127–40.

7. Kringos DS. The strength of primary care in Europe. Dissertation. Utrecht: NIVEL; 2012

8. Kringos DS, Boerma W, van der Zee J, Groenewegen P. Europe’s Strong Primary Care Systems Are Linked To Better Population Health But Also To Higher Health Spending. Health Affairs. 2013;32(4):686–94.

9. Hoffmann K, Ristl R, George A, Maier M, Pichlhöfer O. The ecology of medical care: access points to the health care system in Austria and other developed countries. Scandinavian Journal of Primary Health Care. 2019;37(4):409–17.

10. Hoffmann K, George A, Jirovsky E, Dorner TE. Re-examining access points to the different levels of health care: a cross-sectional series in Austria. European Journal of Public Health. 2019;29(6):1005–10.

11. Hoffmann K, George A, Dorner TE, Süß K, Schäfer WLA, Maier M. Primary health care teams put to the test a cross-sectional study from Austria within the QUALICOPC project. BMC Family Practice. 2015;16(1):168.

12. Main Association of Social Health Insurance Companies in Austria. Teambasierte Primärversorgung 2021. [Team-based Primary Care]. Available from: https://www.sv-primaerversorgung.at/cdscontent/?contentid=10007.796740&viewmode=content [last date accessed 2021-03-09].

13. Amanshauser P. Ideen und Einstellungen der niederösterreichischen Bevölkerung zum Thema Allgemeinmedizin. [Ideas and perception of patients in Lower Austria regarding General Practice]. Diploma thesis. Vienna: Medical University of Vienna; 2020.

14. Förster G. Ideen und Einstellungen zur hausärztlichen Betreuung der Wiener Bevölkerung. [Ideas and perception of patients in Vienna regarding General Practice]. Diploma thesis. Vienna: Medical University of Vienna; 2015.

15. Kaya S. Ideen und Einstellungen der türkischen Bevölkerung zur hausärztlichen Versorgung in Wien. [Ideas and perception of Turkish patients in Vienna regarding General Practice]. Diploma thesis. Vienna: Medical University of Vienna; 2018.

16. Kraml JA. Ideen und Einstellungen der oberösterreichischen Bevölkerung zur hausärztlichen Versorgung. [Ideas and perception of patients in Upper Austria regarding General Practice]. Diploma thesis. Vienna: Medical University of Vienna; 2020.

17. Statista GmbH. Anzahl der Ausländer in Wien nach den zehn wichtigsten Staatsangehörigkeiten zu Jahresbeginn 2021. [Number of foreigners in Vienna at the beginning of the year 2021]. Available from: https://de.statista.com/statistik/daten/studie/886994/umfrage/auslaender-in-wien-nach-staatsangehoerigkeit/ [last date accessed 2021-03-09].

18. Mayring P. Qualitative Inhaltsanalyse. Grundlagen und Techniken. 11 ed. Weinheim: Beltz; 2010.

19. Morgan CL, Beerstecher HJ. Satisfaction, demand, and opening hours in primary care: an observational study. British Journal of General Practice. 2011;61(589):e498–e507.

20. Lippi Bruni M, Mammi I, Ugolini C. Does the extension of primary care practice opening hours reduce the use of emergency services? Journal of Health Economics. 2016;50:144–55.

21. Anderson RT, Camacho FT, Balkrishnan R. Willing to wait?: The influence of patient wait time on satisfaction with primary care. BMC Health Services Research. 2007;7(1):31.

22. Camacho F, Anderson R, Safrit A, Jones AS, Hoffmann P. The Relationship between Patient’s Perceived Waiting Time and Office-Based Practice Satisfaction. North Carolina Medical Journal. 2006;67(6):409–13.

23. Shaw D. The hidden risks of the waiting room: confidentiality and cross-infection. British Journal of General Practice. 2019;69(683):299.

24. Irving G, Neves AL, Dambha-Miller H, Oishi A, Tagashira H, Verho A, et al. International variations in primary care physician consultation time: a systematic review of 67 countries. BMJ Open. 2017;7(10):e017902.

25. Mira JJ, Nebot C, Lorenzo S, Pérez-Jover V. Patient report on information given, consultation time and safety in primary care. Quality and Safety in Health Care. 2010;19(5):e33–e.

26. Ogden J, Bavalia K, Bull M, Frankum S, Goldie C, Gosslau M, et al. “I want more time with my doctor”: a quantitative study of time and the consultation. Family Practice. 2004;21(5):479–83.

27. Dorr Goold S, Lipkin M, Jr. The doctor-patient relationship: challenges, opportunities, and strategies. J Gen Intern Med. 1999;14 Suppl 1(Suppl 1):S26–S33.

28. Kroneman MW, Maarse H, Zee Jvd. Direct access in primary care and patient satisfaction: A European study. Health Policy. 2006;76(1):72–9.

29. EXPH (Expert Panel on Effective Ways of Investing in Health). Report on definition of a frame of reference in relation to primary care with a special emphasis on financing systems and referral systems. Brussels: 2014.

30. World Health Organization. The World Health Report: Primary Health Care—Now More than Ever. Geneva: 2008.

31. O’Malley AS, Tynan A, Cohen GR, Kemper N, Davis MM. Coordination of care by primary care practices: strategies, lessons and implications. Research brief. 2009(12):1–16.

32. Basu S, Phillips RS, Song Z, Bitton A, Landon BE. High Levels Of Capitation Payments Needed To Shift Primary Care Toward Proactive Team And Nonvisit Care. Health Affairs. 2017;36(9):1599–605.

